# Researcher Views on Multi-omics Return of Results to Research Participants: Insights from the Molecular Transducers of Physical Activity Consortium (MoTrPAC) Study

**DOI:** 10.1101/2024.05.15.24307328

**Authors:** Kelly E. Ormond, Caroline Stanclift, Chloe M. Reuter, Jennefer N. Carter, Kathleen E. Murphy, Malene E. Lindholm, Matthew T. Wheeler

## Abstract

**Background:** There is growing consensus in favor of returning individual specific research results that are clinically actionable, valid, and reliable. However, deciding what and how research results should be returned remains a considerable challenge. Researchers are key stakeholders in return of results decision-making and implementation. Multi-omics data contains medically relevant findings that could be considered for return. We sought to understand researchers’ views regarding the potential for multi-omics data derived return of results from a large, national consortium generating multi-omics data.

**Methods:** Researchers from the Molecular Transducers of Physical Activity Consortium (MoTrPAC) were recruited for in-depth semi-structured interviews. To assess understanding of potential clinical utility for types of data collected and attitudes towards return of results in multi-omic clinical studies, we devised an interview guide focusing on types of results generated in the study which could hypothetically be returned based on review of the literature and professional expertise of team members. The semi-structured interviews were recorded, transcribed verbatim and co-coded. Thematic trends were identified for reporting.

**Results:** We interviewed a total of 16 individuals representative of 11 sites and 6 research roles across MoTrPAC. Many respondents expressed positive attitudes regarding hypothetical multi-omics results return, citing participant rights to their data and perception of minimal harm. Ethical and logistical concerns around the return of multi-omics results were raised, including: uncertain clinical validity, a lack of expertise to communicate results, and an unclear obligation regarding whether to return multi-omics results. Further, researchers called for more guidance from funding agencies and increased researcher education regarding return of results.

**Conclusion:** Overall, researchers expressed positive attitudes toward multi-omic return of results in principle, particularly if medically actionable. However, competing ethical considerations, logistical constraints, and need for more external guidance were raised as key implementation concerns. Future studies should consider views and experiences of other relevant stakeholders, specifically clinical genomics professionals and study participants, regarding the clinical utility of multi-omics information and multi-omics results return.

## Background

Human subjects research generates vast amounts of information, some of which holds potential value to study participants (1,2). However the primary goal of human subjects research is to develop results to benefit a greater good. Despite a growing consensus that there is an obligation to return research results to individual study participants that are clinically actionable, valid, and reliable (2–4), deciding what and how results should be returned remains a considerable challenge (5,6). Respect for participants’ autonomy (i.e. their rights to their own data if they so choose), a general moral duty to warn others of imminent harm, and a partial-entrustment account must be weighed against potential harm to study participants and burdens on the research enterprise and on participant-volunteers (6,7). Further, returning research results raises the possibility of therapeutic misconception, when participants confuse the obligations of research versus clinical care (3,8). Therefore, developing return of results protocols for research studies remains an intricate and dynamic problem.

In 2010, an NIH National Heart, Lung, and Blood Institute (NHLBI) expert working group on returning *genomic* individual research results specified that results *should* be offered to study participants who are properly consented and when the following criteria are met; (a) the genetic finding confers <u>substantial risk</u> of health implications (b) there are established therapeutic or preventive <u>interventions</u> that have the potential to change the clinical course of the disease (i.e. are actionable) and (c) the test is <u>analytically valid</u> and disclosure is in accordance with applicable laws (4). This report specified only *genomic* research results, and does not reflect the advancement in high throughput molecular assays and resulting multi-omics research currently being conducted (9). More recently, the National Academies of Sciences, Engineering, and Medicine (NASEM) published a framework (2) to aid in the decision to return individual research results for any tests run on human biospecimens which outlined (1) ‘value to participants’ and (2) ‘feasibility’ should be evaluated on a study-by-study basis. However, what is considered ‘valuable’ to participants is broadly defined in NASEM report. Thus what results researchers are obligated, allowed, or prohibited to return in the current regulatory landscape, particularly those which may not be accurate, clinically actionable, or have clear meaning, remains uncharted territory in practice for multi-omics. Only a few papers have recently begun to help researchers and ethics committees operationalize what the return of individual results processes might look like (for example (10)).

Whereas genomics refers to a comprehensive analysis of genes, multi-omics refers to a global assessment of diverse molecular analytes (11). Multi-omic analyses layer genomics, epigenomics, transcriptomics, proteomics, and metabolomics, often in combination with rich phenotype information, to analyze biological systems, and in many cases to ‘map’ the biology and pathophysiology of human health and disease (11). There are numerous examples showing multi-omics results beyond DNA sequence can contain medically relevant findings (12–16). Plasma untargeted proteomics experiments have demonstrated ability to detect both acute and chronic disease, such as the identification of circulating cardiac troponin in plasma (indicative of myocardial injury) or the quantification of HbA1c, the glycated form of hemoglobin, which when elevated is indicative of diabetes mellitus (12). RNA sequencing (RNA-seq), similar to genomic sequencing, can reliably identify inherited variants which confer an increased risk for diseases with or without established prevention, recommended surveillance, or available treatments (17), while untargeted metabolomics results often contain analytes that are tested in clinical labs to guide clinical care. Additionally, multi-omics results may include data generally recommended not to be disclosed. For example, proteomic analysis of sequence specific peptides of APOE2, APOE3, and APOE4 can provide information regarding risk of Alzheimer’s disease (13). Despite growing evidence that multi-omics results may contain valuable and clinically actionable information for study participants, there is no specific guidance for returning multi-omics results. Further, it is unclear if guidelines for return of genomics results also apply to individual results derived from multi-omics information.

The NIH Common Fund initiative Molecular Transducers of Physical Activity Consortium (MoTrPAC) is an effort across the US to investigate the molecular drivers of exercise adaptation through generation of multi-omic multi-tissue datasets derived from multiple animal and human sub-studies (18–20). The consortium includes multiple clinical research sites, chemical analysis sites, preclinical animal study sites, a bioinformatics center, a clinical coordination center, a data management center and a central biorepository. The clinical sites conduct participant interventions, which include direct interaction with study participants for consent, testing, interventions and biological sampling (blood, lipid, and muscle tissue). The preclinical sites conduct interventions in animal models and perform comprehensive multi-tissue sample collection. The chemical analysis sites are specialized in their particular –omic assays (genomics/epigenomics/transcriptomics, metabolomics or proteomics). The bioinformatics center is tasked with data ingestion, overall quality control, analysis and data dissemination to the consortium and the scientific community. The coordination center, data management center, and biorepository perform central functions which support the operations of the entire consortium (21). MoTrPAC thus includes researchers from many different areas of research expertise, with diverse training, varied experience, and unique perspectives on the study participants, biological sampling, multi-omic data generation and multi-omic results. At the time of this study, there were not yet clear plans to return any MoTrPAC research results directly to individual participants.

Multiple stakeholder groups’ perspectives– including researchers, research participants, health care professionals, Institutional Review Board (IRB) members, sponsors, and the general public– are important to consider in the return of research results process as new technologies arise. Researchers are a particularly critical voice as they are responsible for decisions to return results, protocol development, and implementation (22). To minimize potential burdens of that responsibility and any potential moral distress, their views should be taken into account when developing policy (23). Multiple studies have surveyed researcher views, attitudes, or perspectives about return of results for genomics (eg (24–30). One study found the features of a given condition such as its severity, treatability, and heritability, as well as the perception of clinical validity and certainty of the results are integral factors in the decision to return (25). Dyke et al. (31) examined considerations for return of results in epigenetics research, highlighting among other things the importance of weighing the clinical uncertainty of these results. There is limited literature that has discussed the return of non-genomics omics results. This study seeks to understand researchers’ views regarding the potential for multi-omics return of research results.

## Materials and Methods

### Design

Given the lack of research in this area, we conducted exploratory and in-depth semi-structured interviews with researchers involved with MoTrPAC, a multi-omics research consortium described below, to assess their views regarding the return of multi-omics research results to study participants. IRB approval was received from the Stanford Institutional Review Board (Protocol 57147).

### Subjects and Recruitment

MoTrPAC researchers were invited to participate in semi-structured interviews via a brief recruitment email sent to the consortium. An individual was considered a MoTrPAC researcher if they had an email address affiliated with MoTrPAC, and excluded if they were a member of this study team (MW, ML, KEO). Eligible individuals held a range of roles in the consortium including recruitment or consent responsibilities, exercise physiologists, data production, data analysis, clinical and non-clinical investigators. A link to a Qualtrics survey to screen for inclusion criteria and collect limited demographic information was included in the recruitment email. We employed purposeful sampling to achieve maximum variation amongst interviewees (32). Respondents were stratified based upon demographic information (gender, age, years of experience in human genetics research), role of the researcher in MoTrPAC, and MoTrPAC site.

### Development of the Interview Guide

An interview guide (see Supplemental Methods) was devised from review of literature and professional expertise of team members. Team members held personal experience with offering and returning genetic testing results and multi-omic results (CS, CR, JK, KEO, MW) and had multi-omics and clinical research experience, along with domain expertise in the MoTrPAC human sub-studies (MW, KEO, ML). The interview guide was designed to examine the following domains: (a) understanding of the potential clinical utility for the types of data being collected, (b) knowledge of current plan for return of results in MoTrPAC clinical studies, (c) hypothetical preferences for types of results to be returned in MoTrPAC, and (d) attitudes towards return of multi-omic results. Some education regarding hypothetical types of results which could be returned to participants was provided for all researchers regardless of their prior experience and expertise. For example, actionable genomic results were described as a “change in a gene conferring increased risk for disease that has prevention, recommended surveillance, or available treatments.” When clarification was needed, a specific hypothetical example of a result such as pathogenic variation in *BRCA1* was briefly explained. Researchers were encouraged to ask questions if any concept was unclear to them.

### Interview Process / Data Collection

Semi-structured audio interviews were conducted using the audio-only feature of Zoom Video conferencing platform by a single interviewer (CS) between October 2020 and January 2021. Verbal informed consent was obtained before the interview was conducted. All interviews were audio recorded, transcribed verbatim, de-identified, and checked for accuracy and familiarization.

### Data analysis

We used a thematic analysis approach (33). A codebook was developed by a team (CS, KEM, KEO) based on a combination of literature review and inductive coding from data. This codebook was applied and revised iteratively until consensus was reached after four transcripts. All interview transcripts were then coded by a single researcher (CS) using Dedoose software Version 8.3.45. Every other transcript was co-coded to consensus with a second coder (KEM) to ensure codebook stability and consistency of code application. Coded excerpts were sorted by prevalence and code co-occurrence, then were analyzed for emergent themes. Final themes were agreed upon by consensus of the entire research team. Themes and quotes were selected for inclusion by prevalence or to show breadth of considerations for multi-omics return of results.

## Results

Of the 663 MoTrPAC eligible researchers who received the recruitment email, 35 researchers responded to the enrollment survey (5.3%). Of survey respondents, 31 agreed to be interviewed and 16 completed interviews. Demographics are described in Table 1. The cohort held a range of research functions, including both participant-facing (31%) and non participant-facing (69%) roles, and included Clinical Investigators (12.5%), Scientific Investigators (25%), Recruiter/Consenters (12.5%), Data Producers (12.5%), Data Analysts (25%) and Exercise Physiologists (12.5%). Researchers represented 11 of 31 MoTrPAC sites (35%, not listed due to potential identifiability). Interviews lasted an average of 40 minutes (range 31-57 minutes). Thematic analysis of the data yielded three overarching themes (Table 2): (1) Reasons to support return of individual multi-omics results; (2) Concerns about returning multi-omics results; and (3) The need for guidance regarding return of multi-omic results from funding agencies and national organizations.

**Table 1.** Demographics (N=16)

| **Category** | ***n* (%)** |
| --- | --- |
| **Gender** |  |
| Male | 7 (44%) |
| Female | 9 (56%) |
| **Principal Investigator Status** |  |
| Yes | 6 (37%) |
| No | 10 (63%) |
| **Role within MoTrPaC** |  |
| Recruitment / Consent | 2 (12.5%) |
| Exercise Physiologist | 2 (12.5%) |
| Clinician Investigators (MD, MD/PhD) | 2 (12.5%) |
| Scientist Investigators (PhD) | 4 (25%) |
| Researcher Data Production | 2 (12.5%) |
| Researcher Data Analysis | 4 (25%) |
| **Participant Facing Role** |  |
| Yes | 5 (31%) |
| No | 11 (69%) |
| **Years Research Experience** |  |
| 1-5 | 6 (37%) |
| 6-10 | 4 (25%) |
| >10 | 6 (37%) |

**Table 2.** Thematic analysis yielded 3 overarching themes and relevant sub-themes.

| Themes | Subthemes |
| --- | --- |
| **Theme 1:** Reasons to support return of individual multi-omics results | • *Participant Rights to Their Own Data*<br>• *Perception of Minimal Harms*<br>• *Future Value of Multi-omics Information* |
| **Theme 2:** Concerns about return of multi-omics research results | • *Complex Interpretation with Uncertain Clinical Validity*<br>• *Lack of Clear Obligations to Return Multi-omics Results*<br>• *Expertise Required to Communicate Multi-omics Results* |
| **Theme 3:** Need for external guidance from funding agencies |  |

### Reasons to support return of individual multi-omics results

Reasons given by respondents in support of return of results in general and multi-omics results, specifically, were primarily ethical.

#### Participant Rights to Their Own Data

Researchers expressed high regard for research participants’ autonomy, stressing the importance of informed consent and ability to decide about receiving results based upon participant preferences and values. Twelve of sixteen researchers (12/16, 75%) supported returning multi-omics research information because they believed research participants have a right to the data generated from their individual samples.

Researchers described how participants volunteer “their body, their cells” (Researcher #1, Recruitment/Consent) so data should be returned if they chose. For one researcher, making decisions for patients or participants about which data should be returned was paternalistic:

> “Well you know I don’t like when somebody tries to play the role of God and makes decisions for patients or participants like you know which data should be released and which data should not be released.”
>
> – (Researcher #8, Clinician Investigator)

#### Perception of Minimal Harms

Some researcher’s support for the return of individual multi-omics results relied on the assumption that there is minimal harm in doing so, especially if an expert trained in communicating complicated medical information, such as a genetic counselor, returns the results:

> “I just think all the concern is way overblown and the benefit of providing individuals data about themselves way outweighs the harm… I just don’t see any harms here”
>
> – (Researcher #6, Clinician Investigator)

#### Future Value of Multi-omics Information

Other researchers articulated the potential future value of multi-omics information for research participants’ well-being. They discussed the benefits of providing individuals with information about themselves that they might not learn or have access to outside of a research context, and the constant advancement of scientific knowledge, techniques and analyses. Researchers expected information generated in the study with uncertain clinical validity could yield clinical utility in the future:

> “What we’ve come up with in the last decade and think about what will come up with in the future, in the next decade. I mean, if you could have kind of a catalog of some proteomics or metabolomics information about yourself that at the moment isn’t, or may not be that useful, but just have that in your back pocket and years down the road, be able to plug that into …new analyses and get new information. That would be awesome”
>
> – (Researcher #11, Data Analyst)

Finally, less prominent reasons given in support of returning all research results, including multi-omics, included increased rapport between participants and researchers, promoting altruism, and improved study retention.

> “Rapport, respect, health benefits, health opportunities and just the psychological feeling for the participants that they’ve made a difference, knowing that their results are contributing to the larger picture.”
>
> – (Researcher #4, Exercise Physiologist)

> “We think it’s helpful for retention because anytime you give back to somebody, they’re going to be more favorable toward the study.”
>
> – (Researcher #15, Scientist Investigator)

#### Concerns about returning multi-omics research results

Various ethical and logistical concerns were raised regarding return of results for MoTrPAC in general and for multi-omics results specifically.

#### Complex interpretation with uncertain clinical validity

Researchers stressed the complexities of clinical interpretation of multi-omics data. Some of these concerns were primarily logistical –– they referenced the “sheer volume of data”, potentially time consuming and expensive scientific workflows to find meaning in the data and the ability to execute return of results for a large study cohort;

> “I don’t know what the final sample size will be. But let’s say it’s 2000, around 3000 people. How do we provide results, logistically, to that number of people?… I mean, [sic] highly in favor of giving back results but… I don’t know how they will manage to do that.”
>
> – (Researcher #15, Scientist Investigator)

Others’ concerns were more conceptual, such as the ability to draw accurate conclusions and find clinically actionable information to provide to study participants. The possibility to misinterpret the data led some researchers to suggest multi-omic data could be provided if requested, but without interpretation. While researchers acknowledged the possibility of valuable findings, they stressed the interpretation could be burdensome for the bioinformatics pipeline;

> “I don’t think most of it’s easy to interpret. I think some of it could be… I don’t think most of the information is actionable, quite frankly, unless you go in and spend time on it, meaning you have to do outlier analysis to be able to see if somebody really looks like they have a strange pattern for most of the omics information…but I think on the multi-omics side, it’s possible things will be uncovered and if they’ve consented, yes”
>
> – (Researcher #13, Scientist Investigator)

#### Lack of clear obligation to return multi-omics results

Multiple researchers expressed they did not feel an ethical obligation or moral duty to return multi-omics results due to the perception of unclear clinical validity or utility of these results with current scientific knowledge. A few researchers explicitly mentioned that they felt differently about the obligation to return multi-omic data compared to genomic data, in part because they perceived the health implications are less clear for multi-omics and therefore less concerning. Still others alluded to precedent in the field as justification the obligation to return multi-omics results remains vague;

> “I wouldn’t feel, I wouldn’t have the same kind of moral thoughts of, like, oh, what if we knew something that we didn’t tell them about and then it came back to bite them later? So I don’t know. I could see the proteomics data, like, in particular, what would somebody even do with a bunch of information about their protein expression levels? Is there any sense that they could make out of it? So potentially that information is just not something that’s worth returning raw.”
>
> – (Researcher #11, Data Analyst)

> “I do think that anything where we have clear risk associated with genetic variant, yes, now for all the other classes of data, the proteomics, etc, then unless it points to a genetic variant, the proteomics, metabolomics, the epigenetics all those are usually soft risks. They’re really not established. And nor should I think we worry about those data. Only the genetic data is the domain that I think we need to be concerned about.”
>
> – (Researcher #6, Clinician Investigator)

> “I’m not aware of people that return RNAseq or mass spec back to people like really… I’m not sure we owe it to our participants to give them everything and it’s not typically done.”
>
> – (Researcher #16, Data Analyst)

#### Expertise required to communicate multi-omics results to participants

Finally, many researchers emphasized the need for expertise to translate complex multi-omic data and “package them up in a meaningful way” (Researcher #16, Data Analyst). The ability to answer participant questions about health implications and provide support for follow up care emerged as a key concern. Researchers felt their team members did not have the expertise to answer such questions about multi-omic data. They worried study participants may bring their multi-omics research results to general practitioners unaffiliated with the study, who would likely be unable to answer questions about the meaning of the results or have evidence about the appropriate next steps. Overall, they felt returning results without the ability to accurately answer questions would be futile:

> “What’s going to be returned is something that’s potentially important to their health … Nobody on my team could answer those questions. I don’t think a lot of physicians might necessarily be able to. So I think they’ve got to have the opportunity to go to somebody that could answer any possible questions and there might not be any but I feel very strongly about that because if you just get a piece of paper and you can’t clarify what’s on it by asking somebody it’s almost worthless, frankly.”
>
> – (Researcher #10, Scientist Investigator)

Attitudes about communicating with participants were often shaped by their understanding of clinical testing methods and utility. One researcher felt there is a collective lack of knowledge about potential risks of returning genomics or multi-omics results. They deeply worried about the ability to warn participants of the potential impact receiving results from the study could have;

> “For me as a clinical scientist my primary concern is making sure that I can adequately inform a research volunteer of the risks associated with returning results. And that has not been an easy thing to do for me in MoTrPAC because of the breadth of the type of results that we could ultimately share with participants. It is great from, you know, routine clinical data down to a very deep omics data and helping people understand… we don’t know what the potential clinical impact of that could be and people don’t, most people don’t understand how getting that those results could impact them either favorably or unfavorably so this has been very challenging for me because I don’t know that I feel comfortable in knowing that I can adequately inform participants of what the risks to them might be.”
>
> – (Researcher #9, Scientist Investigator)

Subjective factors, such as personal or family health-related experiences, professional background, and tolerance for uncertainty, often influenced researchers’ views as well. For example, one researcher with clinical training questioned “am I qualified to give that information?… and if so, once they receive that information, handle them from a psychological standpoint?” (Researcher #2, Recruitment/Consent). While this researcher stressed the ability to clearly communicate the information and the psychological impact research results may have, other researchers did not consider psychological ramifications an important factor. One explained “most of the participants that enter into the study to begin with are relatively mentally capable of stepping over minor anxieties so I don’t see that being a contributing challenge” (Researcher #4, Exercise Physiologist). Strikingly, 75% (12/16) of those interviewed thought participants should receive any multi-omics results from a genetic counselor, or at least have access to a genetic counselor. Respondents cited genetic counselors’ expertise in interpreting clinical genomics, delivering complex information, and providing appropriate psychosocial support for clients as part of results disclosure.

In addition to the sub-themes presented above, other concerns raised about return of results in general included participant privacy and potential insurance discrimination. In regard to participant privacy, re-identification was raised as an issue:

> “In this day and age of technology, somebody could decipher your genetic code and figure out that information belongs to you. What are those ramifications and are they prepared for that?”
>
> – (Researcher #2, Recruitment & Consent)

Interestingly, participants only brought up re-identification in the context of genomics results, not multi-omics results which can also be used to re-identify individuals. Still, others worried about insurance discrimination or other financial costs to participants.

#### Need for external guidance from funding agencies and national organizations

In the present study, nearly half of respondents (7/16) specifically mentioned the need for more clear guidance from the NIH and other federal agencies to standardize return of results protocols, particularly for multi-omics projects. Researchers reflected on procedural questions such as funding available within a grant cycle, the need to verify results in CAP accredited and CLIA certified labs, and who would be responsible for facilitating proper referrals or recommendations at the time of disclosure. Many researchers contextualized the problem as “bigger than MoTrPAC”, and cited the growing number of research studies that collect and analyze multi-omic information.

> “So, this is something that should be discussed at a much higher level than just MoTrPAC. I think this should be something that NIH and other federal agencies that are financially supporting these studies, should actually have a policy that should be vetted by medical scientists and clinical researchers.”
>
> – (Researcher #8, Clinician Investigator)

In addition to development of ‘vetted’ policy, researchers felt they lacked knowledge about return of results in general and called for more education on the subject. One researcher brought up the training already required for research personnel and suggested creation of a return of results module:

> “I feel like this is going to be more common practice as we are moving forward in this era of technology. So I think it’s a good thing that we’re looking at this, to figure out, should this be standardized? And, if so, who is standardizing it, you know, across the board? There’s a lot of training as a research professional that you can have, you know, we have your CITI and your Good Clinical Practice. And this may be a whole other module.”
>
> – (Researcher #2, Recruitment/Consent)

These factors and themes map to all steps in the return of results process (Figure 1) including the decision to return results, data generation, data analysis, data validation, communication with participants, and downstream clinical impact.

**Figure 1.**
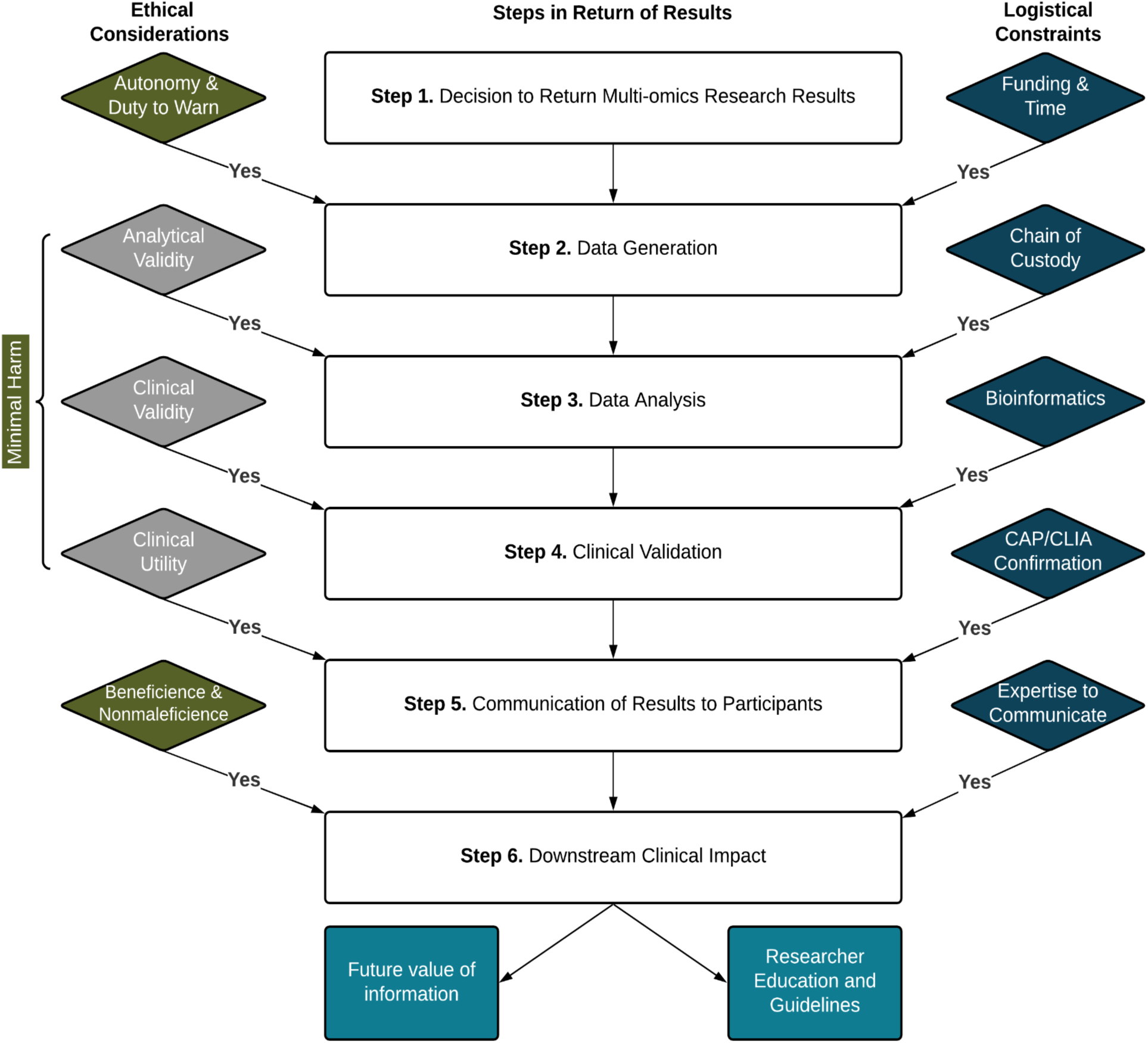
Ethical considerations (left; green), modifiers (left; grey) and logistical constraints (right; blue) from thematic analysis mapped onto steps in the return of results process. Future implications of multi-omics return of results shown at the bottom. Themes were shortened to simplify the diagram.

## Discussion

We conducted semi-structured qualitative interviews to elicit researcher’s views regarding the individual return of multi-omic research results in the context of a specific study, MoTrPAC. Ethical and logistical factors influenced support as well as concerns for return of results. The findings of this study represent unique perspectives about the return of multi-omics research results that have not been described in existing literature.

The majority of researchers in this cohort expressed positive attitudes regarding the return of multi-omics results, particularly if they were considered actionable. This finding is in keeping with researchers’ positive attitudes towards genomics return of results reported previously (23,25,34). Respect for participant autonomy, perception of minimal harms and future value of multi-omics information were brought up as justification for returning multi-omics results, and closely align with the ethical principles outlined by various expert working groups (2,5,35). However, the positive attitudes reported should be interpreted in light of findings from Wynn et al. (36) that suggested researchers who did not have clinical training, provide clinical care to research participants, or have prior experience returning research results were in general more inclined to offer return of results than their colleagues with these characteristics. Furthermore, the depth of the relationship between researcher and participant, degree of dependence or vulnerability of the study population, and the importance of the reasons against return such as costs to the research enterprise or uncertain benefit all have the potential to modify the relative strength of the ethical principles which justify return (5,7).

While researchers in the present study were supportive of returning multi-omics results hypothetically, some questioned whether this was an appropriate use of resources given uncertain clinical validity and utility of data generated in the study. Researchers’ perception of the clinical utility of multi-omics was informed by their knowledge of clinical testing methods and practices as well as understanding of multi-omic data. Given the breadth of information generated in MoTrPAC, researchers stressed the importance of an informed consent process that effectively conveys which results have clearly established clinical utility and which do not. Researchers’ ethical concerns about return of multi-omics results mirror many of the early concerns for genomics return of results (2).

Researchers in this cohort also expressed uncertainty regarding current policy and obligations. Although statements from the NIH specify “as the biomedical research enterprise increasingly moves to a more participatory model of research, where research participants are treated more as partners than passive subjects, we can expect greater emphasis on returning individual-level results of research to participants” (2), researchers felt more specific guidance was required for return of results protocols. These findings suggest there is a need for researcher education on return of results in general and the clinical utility of the multi-omics to ensure this key stakeholder group is able to make evidence-based decisions for implementation.

Practical aspects of return of results such as time, funding and limited access to genetic counselors, are known to influence the decision of researchers or IRB members to return results (25,37). In line with earlier studies (38), researchers in the current work were similarly concerned with the logistical hurdles, particularly for extensive data sets characteristic of multi-omics. Challenges such as extra analyses in the bioinformatics pipeline were often described as burdensome, especially for findings that might not be actionable or align with study aims. Similarly, researchers questioned the infrastructure available to implement a return of results protocol, such as secure portals able to store large multi-omics data sets in a HIPAA compliant manner, funding, genetic counseling resources and follow up care. While logistical barriers and costs are a common theme in return of results literature (2), this work highlights the particular challenges researchers may perceive as barriers in the context of multi-omics ‘big data’.

While feasibility of return is an important question for multi-omics return of results currently, multiple large precision medicine studies have plans or have begun returning medically actionable genetic sequencing results to study participants (39–44). A well known example is the National Institute of Health (NIH)’s All of Us Research Project, which aims to genetically sequence one million people (42). In order to execute returning results, All of Us established a grant funded genetic counseling resource through a private third party company (NIH award OT2 OD028251) which returned information to the first participant in December 2020 and has since returned results to approximately 100,000 participants (45). Other large precision medicine projects such as MyCode at Geisinger Health in collaboration with Regeneron Genetics Center, integrate the return of results process for pathogenic and likely pathogenic variants from a set list of genes found in the research setting into study participants current clinical care by placing the information into their electronic medical record and informing participant’s clinical care team of the research results (40,46,47). Publications detailing the outcomes of these efforts (39–41,48), among others, will undoubtedly help to inform best practices in the future. These works on genomics return of results will also hopefully encourage discussions and engagement from all relevant stakeholders to develop return of results consensus guidelines for other omics in the future.

## Study Limitations and Future Directions

While the researchers interviewed in this study are diverse in specialty, role, institution, and experience and represent views of multi-omic researchers in an NIH funded study distinct from those reported previously in the literature, the majority of participants held non-clinical roles and none of the researchers interviewed were clinical geneticists or genetic counselors. In addition, none of the researchers interviewed had personally returned genomic or multi-omic research results, although two researchers interviewed oversaw protocols where genomic results were returned. Since this study was conducted in 2020-2021, experience and attitudes towards multi-omic data may have changed in the intervening years. Finally, respondents to our recruitment survey may have been biased towards researchers with an interest in or strong opinion on return of results. Future studies should consider views of other relevant stakeholders, specifically study participants, IRBs, and clinical genomics professionals, regarding the return of multi-omics results, and expand the knowledge base regarding the clinical utility of multi-omics information.

## Conclusions

Overall, researchers expressed positive attitudes toward the return of multi-omic research results in principle, citing participant rights to their own data and perception of minimal harm. However, competing ethical considerations, logistical constraints, and a need for more external guidance were raised as key concerns. We provide a roadmap (Figure 1) for future researchers to consider as they design and implement multi-omics return of individual result processes. Future studies also should consider views of other relevant stakeholders, not only study participants, and clinical genomics professionals, but also ethicists, policy makers, legal experts regarding the return of multi-omics research results, and expand the knowledge base regarding the clinical utility of multi-omics information.

## Declarations

### Ethics approval and consent to participate

IRB approval was received from the Stanford Institutional Review Board (Protocol 57147).

### Consent for publication

Not applicable.

### Availability of data and materials

De-identified transcripts, interview guides and other data are available upon reasonable request to the corresponding author.

### Authors’ contributions

**Kelly E. Ormond** – Conceptualization of the project idea; formal analysis; investigation; methodology, supervision, writing-review/editing.

**Caroline Stanclift** – Conceptualization of the project idea; data curation; formal analysis; investigation; methodology; project administration; visualization; writing-original draft preparation.

**Chloe Reuter** – Conceptualization of the project idea; methodology; writing-review/editing.

**Jennefer N. Carter**– Conceptualization of the project idea; methodology; writing-review/editing.

**Kathleen E. Murphy**– Formal analysis; writing-review/editing.

**Malene E. Lindholm**– Conceptualization of the project idea; methodology; recruitment; writing-review/editing.

**Matthew T. Wheeler**– Conceptualization of the project idea; funding acquisition; methodology; supervision; writing-review/editing.

## Acknowledgements

The authors would like to acknowledge Janine Bruce and Sylvia Bereknyei Merrell for their guidance in qualitative research methods. We thank the members of the MoTrPAC Steering Committee for conceptual support and the study personnel who participated in the study. This work was completed in partial fulfillment of the second (CS) author’s degree requirements for the Stanford University Master’s Program in Human Genetics and Genetic Counseling.

## References

1. Wolf SM, Lawrenz FP, Nelson C a., Kahn JP, Cho MK, Clayton W, et al. Managing Incidental Findings in Human Subjects Research: Analysis and Recommendations. J Jaw Med Ethics. 2008;36(2):211–9.

2. National Academies of Sciences, Engineering, and Medicine, Health and Medicine Division, Board on Health Sciences Policy, Committee on the Return of Individual-Specific Research Results Generated, Downey AS, Busta ER, et al. Principles for the Return of Individual Research Results: Ethical and Societal Considerations. National Academies Press (US); 2018.

3. Jarvik GP, Amendola LM, Berg JS, Brothers K, Clayton EW, Chung W, et al. Return of genomic results to research participants: the floor, the ceiling, and the choices in between. Am J Hum Genet. 2014 Jun 5;94(6):818–26.

4. Fabsitz RR, McGuire A, Sharp RR, Puggal M, Beskow LM, Biesecker LG, et al. Ethical and practical guidelines for reporting genetic research results to study participants: updated guidelines from a National Heart, Lung, and Blood Institute working group. Circ Cardiovasc Genet. 2010;3(6):574–80.

5. Belsky L, Richardson HS. Medical researchers’ ancillary clinical care responsibilities. BMJ. 2004 Jun 19;328(7454):1494–6.

6. Berkman BE, Hull SC, Eckstein L. The unintended implications of blurring the line between research and clinical care in a genomic age. Per Med. 2014;11(3):285–95.

7. Richardson HS, Cho MK. Secondary researchers’ duties to return incidental findings and individual research results: a partial-entrustment account. Genet Med. 2012 Apr;14(4):467–72.

8. Lidz CW, Appelbaum PS. The therapeutic misconception: problems and solutions. Med Care. 2002 Sep;40(9 Suppl):V55–63.

9. van Karnebeek CDM, Wortmann SB, Tarailo-Graovac M, Langeveld M, Ferreira CR, van de Kamp JM, et al. The role of the clinician in the multi-omics era: are you ready? J Inherit Metab Dis. 2018 May;41(3):571–82.

10. Waltz A, Johnson B, Schwartz PH. Returning clinically relevant research results to participants: Guidelines for investigators and the IRB. Ethics Hum Res. 2024 Mar;46(2):22–9.

11. Hasin Y, Seldin M, Lusis A. Multi-omics approaches to disease. Genome Biol. 2017 May 5;18(1):83.

12. Chen R, Mias GI, Li-Pook-Than J, Jiang L, Lam HYK, Chen R, et al. Personal omics profiling reveals dynamic molecular and medical phenotypes. Cell. 2012 Mar 16;148(6):1293–307.

13. Geyer PE, Mann SP, Treit PV, Mann M. Plasma Proteomes Can Be Reidentifiable and Potentially Contain Personally Sensitive and Incidental Findings. Mol Cell Proteomics. 2021 Jan 11;20:100035.

14. Yoo BC, Kim KH, Woo SM, Myung JK. Clinical multi-omics strategies for the effective cancer management. J Proteomics. 2018 Sep 30;188:97–106.

15. Kowalczyk T, Ciborowski M, Kisluk J, Kretowski A, Barbas C. Mass spectrometry based proteomics and metabolomics in personalized oncology. Biochim Biophys Acta Mol Basis Dis. 2020 May 1;1866(5):165690.

16. Özdemir V, Dove ES, Gürsoy UK, Şardaş S, Yıldırım A, Yılmaz ŞG, et al. Personalized medicine beyond genomics: alternative futures in big data— proteomics, environtome and the social proteome. J Neural Transm. 2017 Jan 1;124(1):25–32.

17. Piskol R, Ramaswami G, Li JB. Reliable identification of genomic variants from RNA-seq data. Am J Hum Genet. 2013 Oct 3;93(4):641–51.

18. Amar D, Gay NR, Jimenez-Morales D, Jean Beltran PM, Ramaker ME, Raja AN, et al. The mitochondrial multi-omic response to exercise training across rat tissues. Cell Metab [Internet]. 2024 Apr 15; Available from: 10.1016/j.cmet.2023.12.021

19. MoTrPAC Study Group, Lead Analysts, MoTrPAC Study Group. Temporal dynamics of the multi-omic response to endurance exercise training. Nature. 2024 May;629(8010):174–83.

20. Study Group, MoTrPAC. Molecular Transducers of Physical Activity Consortium (MoTrPAC): Human Studies Design and Protocol. J Appl Physiol [Internet]. 2024 Apr 18; Available from: 10.1152/japplphysiol.00102.2024

21. Sanford JA, Nogiec CD, Lindholm ME, Adkins JN, Amar D, Dasari S, et al. Molecular Transducers of Physical Activity Consortium (MoTrPAC): Mapping the Dynamic Responses to Exercise. Cell. 2020 Jun 25;181(7):1464–74.

22. Kinsella K. Issues in Returning Individual Results from Genome Research Using Population-based Banked Specimens, with a Focus on the National Health and Nutrition Examination Survey: Workshop Summary. National Academies Press; 2014. 95 p.

23. Richter G, De Clercq E, Mertz M, Buyx A. Chapter 6 – Reporting of secondary findings in genomic research: Stakeholders’ attitudes and preferences. In: Langanke M, Erdmann† P, Brothers KB, editors. Secondary Findings in Genomic Research. Academic Press; 2020. p. 99–132.

24. Fernandez CV, Strahlendorf C, Avard D, Knoppers BM, O’Connell C, Bouffet E, et al. Attitudes of Canadian researchers toward the return to participants of incidental and targeted genomic findings obtained in a pediatric research setting. Genet Med. 2013 Jul;15(7):558–64.

25. Klitzman R, Appelbaum PS, Fyer A, Martinez J, Buquez B, Wynn J, et al. Researchers’ views on return of incidental genomic research results: qualitative and quantitative findings. Genet Med. 2013 Nov;15(11):888–95.

26. Kostick KM, Brannan C, Pereira S, Lázaro-Muñoz G. Psychiatric genetics researchers’ views on offering return of results to individual participants. Am J Med Genet B Neuropsychiatr Genet. 2019 Dec;180(8):589–600.

27. Lázaro-Muñoz G, Torgerson L, Pereira S. Return of results in a global survey of psychiatric genetics researchers: practices, attitudes, and knowledge. Genet Med. 2021 Feb;23(2):298–305.

28. Middleton A, Morley KI, Bragin E, Firth HV, Hurles ME, Wright CF, et al. Attitudes of nearly 7000 health professionals, genomic researchers and publics toward the return of incidental results from sequencing research. Eur J Hum Genet. 2016 Jan;24(1):21–9.

29. Mwaka ES, Sebatta DE, Ochieng J, Munabi IG, Bagenda G, Ainembabazi D, et al. Researchers’ perspectives on return of individual genetics results to research participants: a qualitative study. Global Bioethics. 2021 Jan 1;32(1):15–33.

30. Ramoni RB, McGuire AL, Robinson JO, Morley DS, Plon SE, Joffe S. Experiences and attitudes of genome investigators regarding return of individual genetic test results. Genet Med. 2013 Nov;15(11):882–7.

31. Dyke SOM, Saulnier KM, Dupras C, Webster AP, Maschke K, Rothstein M, et al. Points-to-consider on the return of results in epigenetic research. Genome Med. 2019 May 23;11(1):31.

32. Lawrence A. Palinkas • Sarah M. Horwitz •, Carla A. Green • Jennifer P. Wisdom •, Naihua Duan • Kimberly Hoagwood. Purposeful Sampling for Qualitative Data Collection and Analysis in Mixed Method Implementation Research. Adm Policy Ment Health. 2015;:533–544:533–44.

33. Chapman AL, Hadfield M, Chapman CJ. Qualitative research in healthcare: an introduction to grounded theory using thematic analysis. J R Coll Physicians Edinb. 2015;45(3):201–5.

34. Scherr CL, Aufox S, Ross AA, Ramesh S, Wicklund CA, Smith M. What People Want to Know About Their Genes: A Critical Review of the Literature on Large-Scale Genome Sequencing Studies. Healthcare (Basel) [Internet]. 2018 Aug 8;6(3). Available from: 10.3390/healthcare6030096

35. Fabitz RR, McGuire A, Sharp R, Puggal M, Beskow LM. Ethical and practical guidelines for reporting genetic research results to study participants. Circ Cardiovasc Genet.

36. Wynn J, Martinez J, Duong J, Zhang Y, Phelan J, Fyer A, et al. Association of Researcher Characteristics with Views on Return of Incidental Findings from Genomic Research. J Genet Couns. 2015 Oct;24(5):833–41.

37. Keogh LA, Fisher D, Sheinfeld Gorin S, Schully SD, Lowery JT, Ahnen DJ, et al. How do researchers manage genetic results in practice? The experience of the multinational Colon Cancer Family Registry. J Community Genet. 2014 Apr;5(2):99–108.

38. Meacham MC, Starks H, Burke W, Edwards K. Researcher perspectives on disclosure of incidental findings in genetic research. J Empir Res Hum Res Ethics. 2010 Sep;5(3):31–41.

39. Rego S, Dagan-Rosenfeld O, Bivona SA, Snyder MP, Ormond KE. Much ado about nothing: A qualitative study of the experiences of an average-risk population receiving results of exome sequencing. J Genet Couns [Internet]. 2019 Mar 5; Available from: 10.1002/jgc4.1096

40. Schwartz MLB, McCormick CZ, Lazzeri AL, Lindbuchler DM, Hallquist MLG, Manickam K, et al. A Model for Genome-First Care: Returning Secondary Genomic Findings to Participants and Their Healthcare Providers in a Large Research Cohort. Am J Hum Genet. 2018 Sep 6;103(3):328–37.

41. Suckiel SA, O’Daniel JM, Donohue KE, Gallagher KM, Gilmore MJ, Hendon LG, et al. Genomic Sequencing Results Disclosure in Diverse and Medically Underserved Populations: Themes, Challenges, and Strategies from the CSER Consortium. J Pers Med [Internet]. 2021 Mar 13;11(3). Available from: 10.3390/jpm11030202

42. All of Us Research Program | NIH [Internet]. 2020 [cited 2024 May 9]. All of Us Research Program. Available from: https://allofus.nih.gov/

43. UK Biobank. UK Biobank ethics and governance framework version 3.0. 2007;0(July):20.

44. Leppig KA, Kulchak Rahm A, Appelbaum P, Aufox S, Bland HT, Buchanan A, et al. The reckoning: The return of genomic results to 1444 participants across the eMERGE3 Network. Genet Med. 2022 May;24(5):1130–8.

45. All of Us Research Program | NIH [Internet]. 2024 [cited 2024 May 9]. All of Us Returns Health-Related DNA Results to 100,000 Participants. Available from: https://allofus.nih.gov/news-events/announcements/all-us-returns-health-related-dna-results-100000-participants

46. Carey DJ, Fetterolf SN, Davis FD, Faucett WA, Kirchner HL, Mirshahi U, et al. The Geisinger MyCode community health initiative: an electronic health record-linked biobank for precision medicine research. Genet Med. 2016 Sep;18(9):906–13.

47. Faucett WA, Davis FD. How Geisinger made the case for an institutional duty to return genomic results to biobank participants. Appl Transl Genom. 2016 Mar;8:33–5.

48. Berg JS, Amendola LM, Eng C, Van Allen E, Gray SW, Wagle N, et al. Processes and preliminary outputs for identification of actionable genes as incidental findings in genomic sequence data in the Clinical Sequencing Exploratory Research Consortium. Genet Med. 2013 Nov;15(11):860–7.

